## Supplementary material for "Common misconceptions held by health researchers when interpreting linear regression assumptions, a cross-sectional study": S1 Table: Formatted linear regression statistical questionnaire.

The answers to this survey should be only based on the linear regression aspects of the papers. For the purpose of this study, please also exclude analyses which are identified as ANOVA, have accounted for clustering or random effects, non-parametric linear regression models, Bayesian or other alternative methods to linear regression Note: general linear models, ANCOVA and generalised linear models where they refer to linear regression should be included.

(Q1) Please enter the full name of the first author as it appears on the paper, including first name, surname

(Q2) Do you have a conflict of interest in reviewing this paper?

- ○ No (go to next question)
- ○ Yes (if yes, do not continue)

How many linear regressions can you identify in this paper? Include as a separate regression those using a different dependent variable, different independent variable(s) or different data. For example, a paper with five univariate results plus one multivariable model should be counted as six regressions. Analyses should be counted if any result considered to be a linear regression is presented, eg., R 2 , p-value, etc.

(Q3) Number of linear regressions (if the answer is 0 linear regressions do not continue with question)

(3a) Any comments

(Q4) Does the paper mention a strategy for assessing linear regression assumptions?

- ○ No
- ○ Yes

The assumptions below should only be answered considering the linear regression models in the paper.

(Q5) Did the authors check the normality assumption?

- ○ No (go to Q8)
- ○ Yes (answer questions Q6 and Q7)

(Q6) What was checked with regards to normality? (tick as many that apply)

- ☐ Unclear
- ☐ Y variable
- ☐ X variable
- ☐ Sub groups of Y
- ☐ Residuals

Other (please specify)

(Q7) How was normality assessed? (tick as many that apply)

- ☐ Unclear
- ☐ Not described
- ☐ Descriptive statistics (skewness, kurtosis etc.)
- ☐ Plots (including residual plots)
- ☐ Statistical test

Other (please specify)

(Q8) Did the authors check the linearity assumption?

- ○ No (go to Q10)
- ○ Yes (answer Q9)

(Q9) How was linearity assessed? (tick as many that apply)

- ☐ Unclear
- ☐ Not described
- ☐ Raw data
- ☐ Plots (including residual plots)
- ☐ Residuals
- ☐ Statistical test

Other (please specify)

(Q10) Did the authors check the homoscedasticity?

- ○ No (go to Q12)
- ○ Yes (answer Q11)

(Q11) How was homoscedasticity assessed? (tick as many that apply)

- ☐ Unclear
- ☐ Not described
- ☐ Raw data
- ☐ Plots (including residual plots)
- ☐ Residuals
- ☐ Statistical test

Other (please specify)

(Q12) Did the authors check the independence of observations?

- ○ No (go to Q14)
- ○ Yes (answer Q13)

(Q13) How was independence of observations assessed? (tick as many that apply)

- ☐ Unclear
- ☐ Not described
- ☐ Authors stated independent design
- ☐ Raw data
- ☐ Plots (including residual plots)
- ☐ Residuals
- ☐ Statistical test

Other (please specify)

(Q14) Was collinearity of X variables in models evaluated?

- ○ Not required
- ○ No
- ○ Yes

(Q15) Have authors checked for outliers in their data?

- ○ No (go to Q17)
- ○ Yes (answer Q16)

(Q16) What did they do with respect to outliers?

- Unclear
- No action taken
- Outliers removed from all analyses
- Sensitivity analysis
- Data transformation
- Bootstrapped

Other (please specify)

(Q17) Were any continuous variables transformed, not including categorisation?

- No
- Yes, but did not describe reasoning for transformation
- Yes, described reasoning for transformation

(Q18) Were continuous variables on a very large/small scales in the model scaled appropriately?

- Not required
- Unclear
- No
- Yes

(Q19) Is there any process for selecting the variables included in the final model? (only choose one)

- Unclear (go onto Q22)
- Univariate modeling only (one X variable) (go on to Q22)
- Model based on reference literature or author knowledge (go to Q22)
- Significant variables from univariate analysis were included in a multivariable model (go to Q22)
- Used recognised statistical modeling strategy (go Q20 and Q21)

Other (please specify) (go to Q22)

(Q20) Which variable selection strategy was used? (only choose one)

- ○ Forwards
- ○ Backwards
- ○ Stepwise
- ○ Information criterion/theory (AIC/BIC, etc.)
- ○ Regularisation methods (elasticnet, ridge regression, lasso etc.)

Other (please specify)

(Q21) Does the paper mention any statistical significance criteria for including variables?

- ○ No
- ○ Yes

(Q22) What has been reported? (Tick all the apply)

If coefficients not reported skip Q23, 24,and Q25

- ☐ Coefficients
- ☐ Confidence intervals
- ☐ Standard error
- ☐ R-Squared
- ☐ F/t statistics
- ☐ Degrees of freedom
- ☐ Number of observations in models

(Q23) Has the direction of the parameter estimates been interpreted? (if coefficients present)

- ○ No
- ○ Yes

(Q24) Has the size of the parameter estimates been interpreted? (if coefficients present)

- ○ No
- ○ Yes

(Q25) Have authors discussed the scientific importance of parameters estimates? (if coefficients present)

- ○ No
- ○ Yes, but only in a generic way
- ○ Yes, linked size of effect back to outcome variable

(Q26) Have p-values been reported?

- ○ No
- ○ Mostly reported categorically eg <0.05, <0.01
- ○ Mostly reported continuously

(Q27) Was linear regression the main analysis in the paper?

- ○ No
- ○ Yes

(Q28) Was most of the detail for the assumption checks in the supporting information (appendix)?

- ○ No
- ○ Yes

(Q29) In terms of the linear regression analyses, rate the statistical quality of the

| | Very poor | Poor | Fair | Good | Very good |
|---|---|---|---|---|---|
| | ○ | ○ | ○ | ○ | ○ |

(Q30) Do you have any other comments you would like to provide with respect to the linear regression and their quality in the paper?

Any more comments?
