## Supplementary material for "Common misconceptions held by health researchers when interpreting linear regression assumptions, a cross-sectional study": S3 Table: Full reporting of agreement and reliability for statistical raters.

| Variable | Agreement | Pe | Gwet | 95% CI | SE | p-value |
| --- | --- | --- | --- | --- | --- | --- |
| **Rating 1 vs Rating 2** |  |  |  |  |  |  |
| Normality | 88% | 38% | 0.81 | 0.70, 0.93 | 0.06 | <0.001 |
| Linearity | 89% | 26% | 0.85 | 0.75, 0.96 | 0.05 | <0.001 |
| Homoscedasticity | 100% |  |  |  |  |  |
| Independence | 78% | 29% | 0.69 | 0.55, 0.83 | 0.07 | <0.001 |
| Outliers | 83% | 4% | 0.82 | 0.74, 0.91 | 0.04 | <0.001 |
| Rating | 91% | 67% | 0.74 | 0.64, 0.83 | 0.05 | <0.001 |
| **Rating 1 vs Prevalence** |  |  |  |  |  |  |
| Normality | 91% | 40% | 0.84 | 0.74, 0.95 | 0.05 | <0.001 |
| Linearity | 92% | 27% | 0.88 | 0.79, 0.97 | 0.05 | <0.001 |
| Homoscedasticity | 98% | 10% | 0.98 | 0.94, 1.00 | 0.02 | <0.001 |
| Independence | 91% | 16% | 0.89 | 0.81, 0.96 | 0.04 | <0.001 |
| Outliers | 91% | 5% | 0.90 | 0.84, 0.96 | 0.03 | <0.001 |
| **Rating 2 vs Prevalence** |  |  |  |  |  |  |
| Normality | 92% | 40% | 0.86 | 0.76, 0.96 | 0.05 | <0.001 |
| Linearity | 90% | 28% | 0.87 | 0.77, 0.96 | 0.05 | <0.001 |
| Homoscedasticity | 98% | 10% | 0.98 | 0.94, 1.00 | 0.02 | <0.001 |
| Independence | 83% | 24% | 0.78 | 0.66, 0.90 | 0.06 | <0.001 |
| Outliers | 86% | 5% | 0.86 | 0.78, 0.93 | 0.04 | <0.001 |

Agreement = Observed agreement, Pe = The expected agreement by chance, Gwet = Gwet agreement coefficient, 95% CI = Gwet 95% confidence intervals, SE = Standard Error. Homoscedasticity had 100% agreement for the two raters resulting in a Gwet SE of zero, therefore, these results were not reported. The paper rating was only of interest between the two raters, and was scored on a Likert scale, and was analysed using quadratic weights; all other variables were binary and did not require weighting.
